# National genomic profiling of *Plasmodium falciparum* antimalarial resistance in Zambian children participating in the 2018 Malaria Indicator Survey

**DOI:** 10.1101/2024.08.05.24311512

**Authors:** Abebe A. Fola, Ilinca I. Ciubotariu, Jack Dorman, Mulenga C. Mwenda, Brenda Mambwe, Conceptor Mulube, Rachael Kasaro, Moonga B. Hawela, Busiku Hamainza, John M. Miller, Jeffrey A. Bailey, William J. Moss, Daniel J. Bridges, Giovanna Carpi

## Abstract

The emergence of antimalarial drug resistance is a major threat to malaria control and elimination. Using whole genome sequencing of 282 *P. falciparum* samples collected during the 2018 Zambia National Malaria Indicator Survey, we determined the prevalence and spatial distribution of known and candidate antimalarial drug resistance mutations. High levels of genotypic resistance were found across Zambia to pyrimethamine, with over 94% (n=266) of samples having the *Pfdhfr* triple mutant (N51**I**, C59**R**, and S108**N**), and sulfadoxine, with over 84% (n=238) having the *Pfdhps* double mutant (A437**G** and K540**E**). In northern Zambia, 5.3% (n=15) of samples also harbored the *Pfdhps* A581**G** mutation. Although 29 mutations were identified in *Pfkelch13*, these mutations were present at low frequency (<2.5%), and only three were WHO-validated artemisinin partial resistance mutations: P441**L** (n=1, 0.35%), V568**M** (n=2, 0.7%) and R622**T** (n=1, 0.35%). Notably, 91 (32%) of samples carried the E431**K** mutation in the *Pfatpase6* gene, which is associated with artemisinin resistance. No specimens carried any known mutations associated with chloroquine resistance in the *Pfcrt* gene (codons 72-76). *P. falciparum* strains circulating in Zambia were highly resistant to sulfadoxine and pyrimethamine but remained susceptible to chloroquine and artemisinin. Despite this encouraging finding, early genetic signs of developing artemisinin resistance highlight the urgent need for continued vigilance and expanded routine genomic surveillance to monitor these changes.

## Introduction

Among all *Plasmodium* species that infect humans, *P. falciparum* is of the greatest significance, accounting for over 95% of malaria deaths.^1,2^ The World Health Organization (WHO) recommends front line artemisinin combination therapy (ACT), such as artemether-lumefantrine (AL), artesunate-amodiaquine (ASAQ) or dihydroartemisinin-piperaquine (DHAP), in most African countries for treatment of malaria.^3^ For prevention, sulfadoxine-pyrimethamine (SP) is recommended for intermittent preventive treatment in pregnancy (IPTp) and for infants (IPTi) living in high-transmission areas.^3,4^ Antimalarial drugs have played a major role in achieving a significant reduction in the malaria burden globally since 2000 but progress towards malaria elimination has stalled, with resurgence in several endemic countries.^5,6^ A major threat to control and elimination is the emergence and spread of antimalarial drug resistance.

*P. falciparum* parasites have developed resistance to nearly every available antimalarial drug and resistant strains have spread across malaria endemic countries.^7^ For example, *P. falciparum* resistance to chloroquine (CQ) emerged in 1957 in Thailand, spread to Southeast Asia in the 1970’s, and by 1982 resistance had spread across the entire African continent.^8^ Several alleles are associated with CQ resistance around codon 72-76 in the *P. falciparum* CQ resistance transporter gene (*Pfcrt*), including the diagnostic K76**T** mutation.^9^ Similarly, mutations in the enzymes dihydropteroate synthase (*Pfdhps*) (S436**A**, A437**G**, K540**E**, A581**G**, A613**S**) and dihydrofolate reductase (*Pfdhfr*) (N51**I**, C59**R**, S108**N**, I164**L**) are associated with varying degree of resistance to sulfadoxine and pyrimethamine, respectively,^10,11^ and are widespread in Africa, Asia, South America, and Oceania.^12^ Combinations of these mutations i.e., triple *Pfdhfr* mutations of N51**I**, C59**R**, and S108**N**, plus double *Pfdhps* mutations of A437**G**, K540**E** (IRNGE - ‘Quintuple mutant’) confer full resistance to SP.^4,13^ Moreover, parasites that have the additional *Pfdhps* A581**G** mutation (IRNGEG - ‘Sextuple-mutant’) are associated with enhanced SP resistance *in vitro* that contribute to super SP resistance and IPTp failure.^14,15^

Since 2008, *P. falciparum* parasites resistant to first-line artemisinin (ART) treatments have emerged in Southeast Asia^16,17^ and spread in the Greater Mekong Subregion (GMS).^18^ Studies identified point mutations in the beta-propeller domain of *kelch* 13 (*Pfkelch13*) that were associated with reduced susceptibility to ART and its derivatives, manifested by delayed parasite clearance times.^19^ To classify ART resistant (ART-R) parasites, WHO provided a list of 9 “validated” and 12 “associated/candidate” *kelch13* ART resistance markers.^20^ Resistance to partner drugs has also been identified, specifically mutations N86**Y**, Y184**F** and D1246**Y** in the *P. falciparum* multidrug resistance gene 1 (*Pfmdr1*), which have been associated with reduced susceptibility to lumefantrine.^21^ With reports of decreased ACT efficacy and treatment failure in Africa,^22–24^ and genotypic identification of an increasing prevalence of WHO-validated *kelch13* ART-R marker (R561**H**),^25,26^ the threat of antimalarial resistance is increasing such that surveillance should be high priority.^27,28^

In Zambia, despite the continued use of SP for IPTp and AL for the treatment of uncomplicated malaria since 2002 (the first African country to adopt AL as a first-line treatment policy nationwide), only a few small-scale studies in Southern, Western, and Luapula Provinces have investigated the prevalence of CQ and ACT resistance markers.^29,30^ However, our recent nationwide genomic study,^31^ aimed at understanding malaria transmission across Zambia and establishing a baseline for parasite genetic metrics, identified strong positive selection signatures in genes involved in SP and ACT resistance that warrants further investigation into the regional prevalence and spatial variations of drug resistance markers. Moreover, genetic background mutations that could augment ART-R and other novel mutations that may confer antimalarial resistance have yet to be thoroughly assessed in Zambia.

To address these important knowledge gaps and support the Zambian National Malaria Elimination Program, we conducted a genomic surveillance study nested within the 2018 Zambia Malaria Indicator Survey (MIS), which comprises nationwide representative samples.^31^ We generated 282 *P. falciparum* whole genome sequences (WGS) from seven provinces and mined these data for known and candidate antimalarial resistance mutations across 5 key *P. falciparum* drug resistance genes *(Pfdhfr*, *Pfdhps*, *Pfkelch13*, *Pfcrt*, and *Pfmdr1*) and other gene that may contribute to ART-R phenotypes: apicoplast ribosomal protein S10 precursor *(Pfarps10),* multidrug resistance protein 2 *(Pfmdr2),* ferredoxin (*Pffd),* adaptor protein 2 complex subunit mu gene (*Pfap2mu*), ubiquitin carboxyl-terminal hydrolase 1 gene (*Pfubp1*),^32,33^ and reticulum Ca^2+^ ATPase (*Pfatp6*).^34^ This study defines the most complete genetic landscape of antimalarial drug resistance markers in Zambia, allows spatial and temporal trends to b identified, and provides the foundation for future studies.

## Results

### Whole genome sequencing and mining of drug resistance markers

A total of 282 specimens collected during the 2018 Zambia MIS from children younger than five years of age (**Figure 1**), and were whole genome sequenced (**Figure S1**) with high coverage across 13 *P. falciparum* genes associated with antimalarial drug resistance (**Table S1**). Within the open-reading frames of these genes, 489 non-synonymous (NS) mutations (**Table S2** and **Figure S2**) were identified with variable spatial frequencies across Zambia (**Table S3**). The prevalence of key mutations associated with antimalarial drug resistance is shown in **Table 1**.

**Figure 1:**
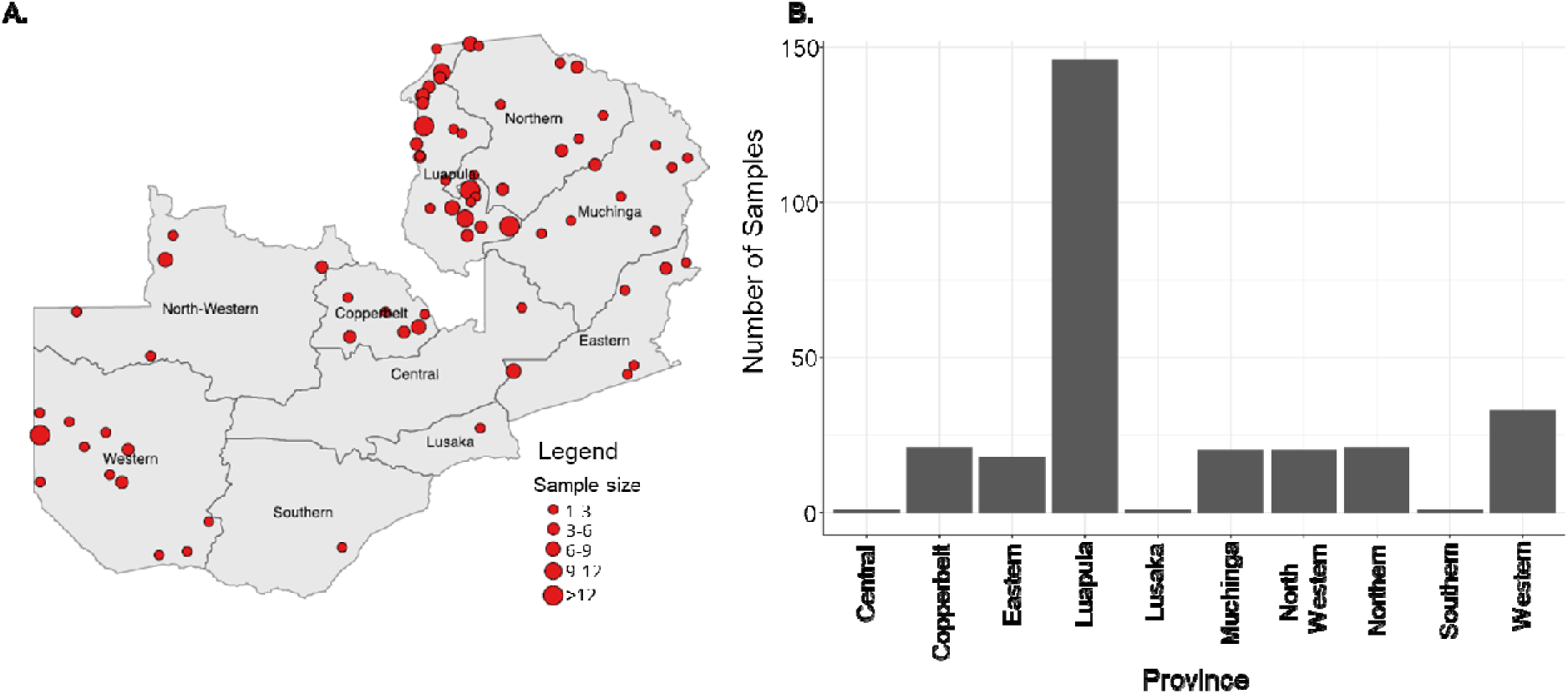
Spatial distribution of samples retained (n=282) for downstream drug resistanc analysis. **A)** Sample distribution at the cluster level across Zambia. The size of each sample collection cluster (red) is shown in proportion to the cluster sample size. **B)** Sample size per province. Three provinces (Central, Lusaka and Southern) were excluded from provincial prevalence calculations due to low sample size.

**Table 1:**
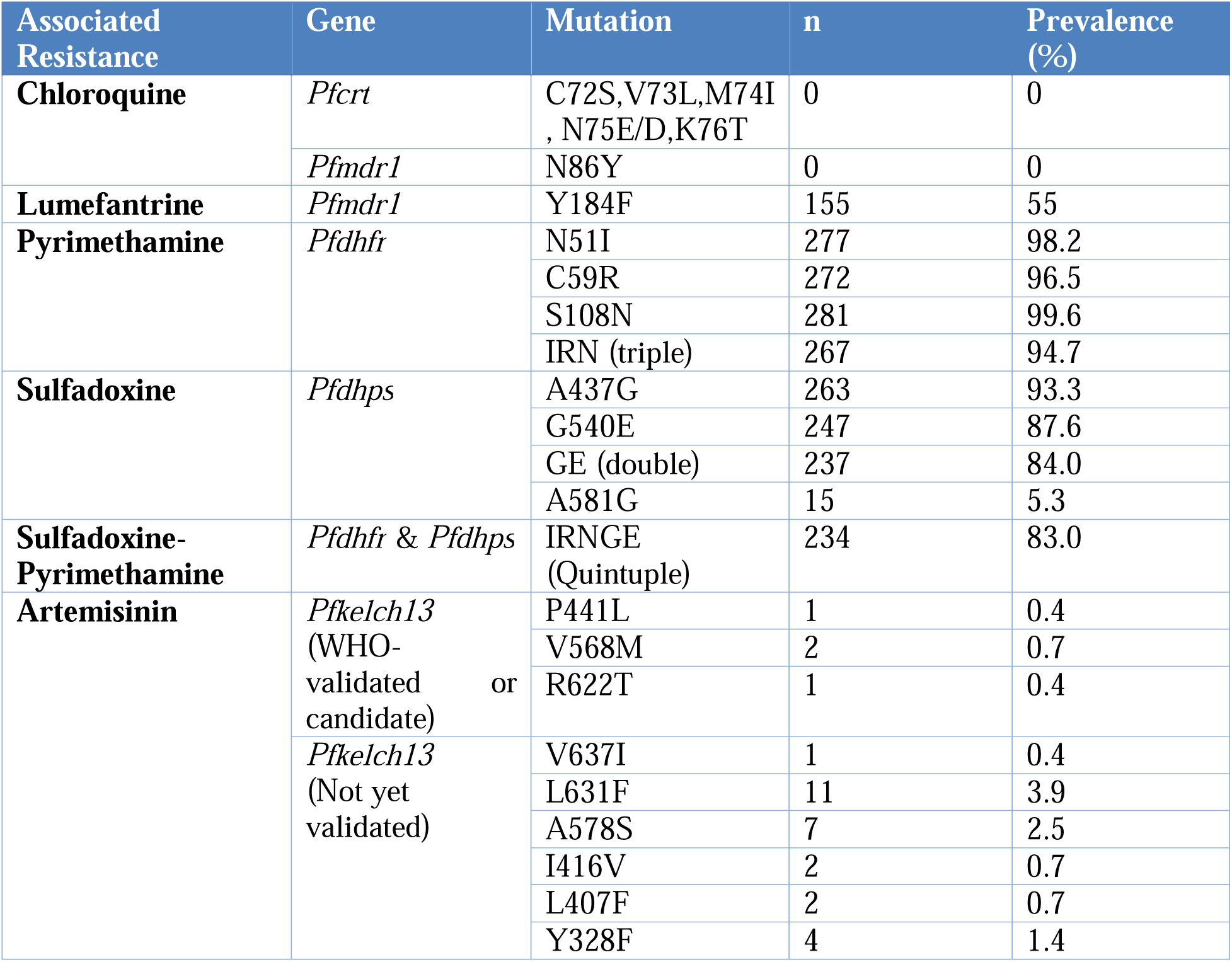
Prevalence of key mutations associated with mono- and multiple antimalarial drug resistance across Zambia in 2018. n = number of samples that carried the mutant allele out of the total sequenced samples (282)

### Chloroquine resistance

The *Pfcrt* K76**T** mutation associated with CQ resistance was not identified in the analyzed samples (**Table 1, Table S2**), indicating reversal of CQ sensitive *P. falciparum* strains across Zambia.^31,35^ Seven NS mutations not associated with CQ resistance were identified at very low frequencies in other codons across the *Pfcrt* gene (PF3D7_0709000), but 94.0% (265/282) of sequenced samples did not carry any of these mutations (**Figure S3, Table S2**). All sequenced specimens were wild-type (N) at codon 86 in the multidrug resistance transporter *Pfmdr1* gene, which has been shown to enhance resistance to CQ in some genetic backgrounds,^36^ further supporting reversal to CQ sensitive parasites in Zambia.

### Antifolate resistance

Key mutations (N51**I**, C59**R**, S108**N**) in the *Pfdhfr* gene were identified in 98.2% (277/282), 96.5% (272/282), and 99.6% (281/282) of samples, respectively, with 94.6% (267/282) classified as a *Pfdhfr* triple (**IRN**) mutants (**Table 1 and Figure 2A**). Moreover, key mutations A437**G** and K540**E**, in the *Pfdhps* gene were identified in 93.3% (263/282) and 87.6% (247/282) of sequenced samples, respectively, with 84.0% (237/282) classified as a *Pfdhps* double (**GE**) mutant (**Table 1 and Figure 2C**). We found some degree of spatial heterogeneity for both *Pfdhfr* triple mutants and *Pfdhps* double mutants at the cluster level across the country (**Figure 2B** and **Figure 2D**, respectively).

**Figure 2.**
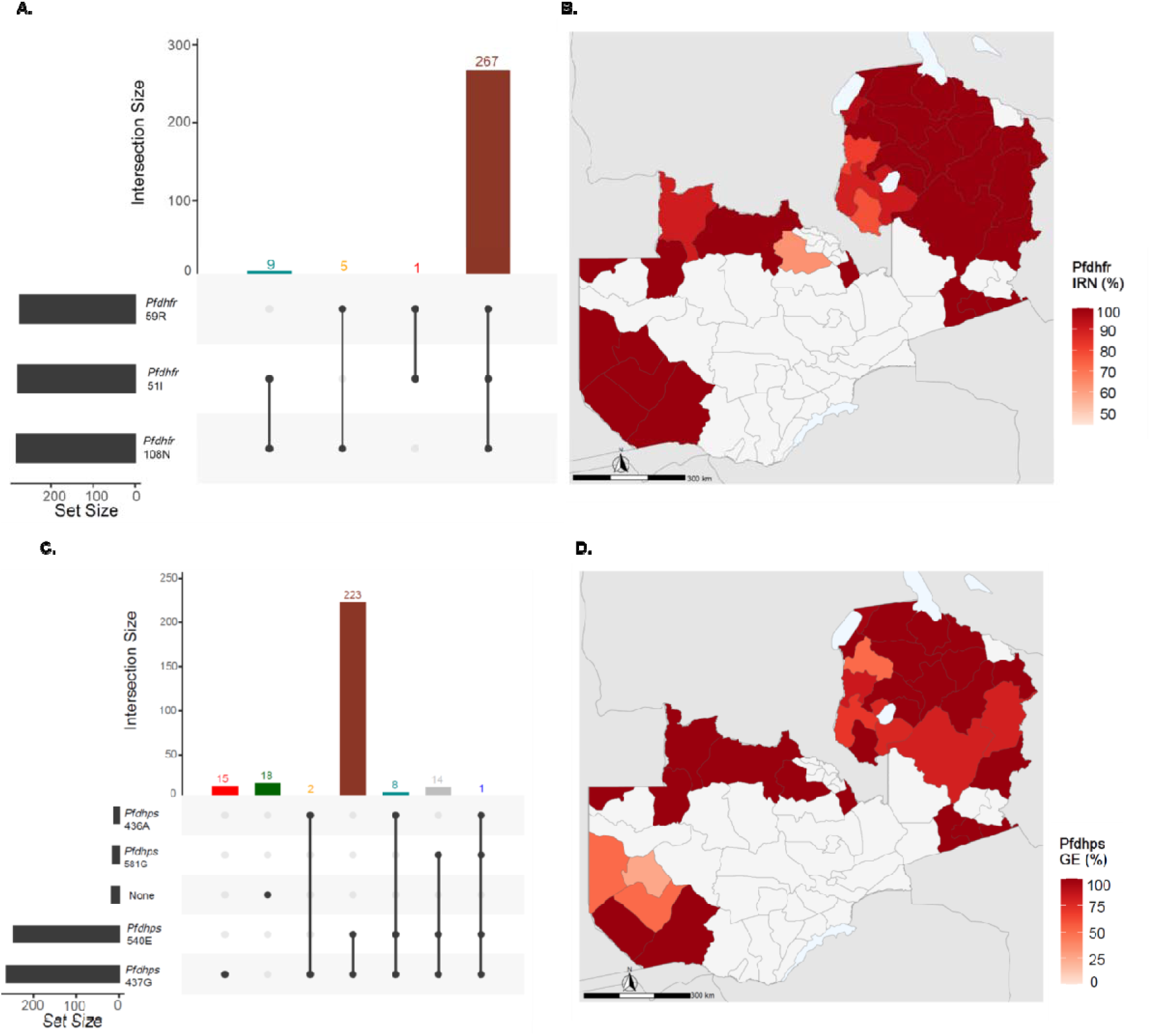
Prevalence of *Pfdhfr* and *Pfdhps* mutations across Zambia. UpSet plots showing the number of times each combination of mutations was seen for *Pfdhfr* (**A**) and *Pfdhps* (**C**). The lines and circles below the bars represent the different combinations of resistant genotype in individual samples. **B**) Spatial prevalence of pyrimethamine resistant triple mutant (*Pfdhf* 51**I***/*59**R**/108**N**, **IRN**), **D**) Spatial prevalence of sulfadoxine resistant double mutant (*Pfdhp* 437**G***/*540**E**, **GE**). Color code heat map shows prevalence at the district level, while white indicates districts where no samples were available.

Overall, 83.0% (234/282) of sequenced samples were typed as quintuple mutants (*Pfdhfr* IRN, *Pfdhps* GE, **Figure 3A**), a genotype associated with near complete SP resistance (**Table 1**). In the *Pfdhps* gene, the A581**G** mutation was found in 5.3% (15/282) of the analyzed samples with high geographic variation across Zambia and with clustering in the adjacent Luapula and Northern Provinces (**Figure 3B**). There was no evidence for the presence of *Pfdhfr* I164**L** or *Pfdhps* A613**S**/**T**, mutations that can enhance the quintuple mutant SP resistance profile.

**Figure 3.**
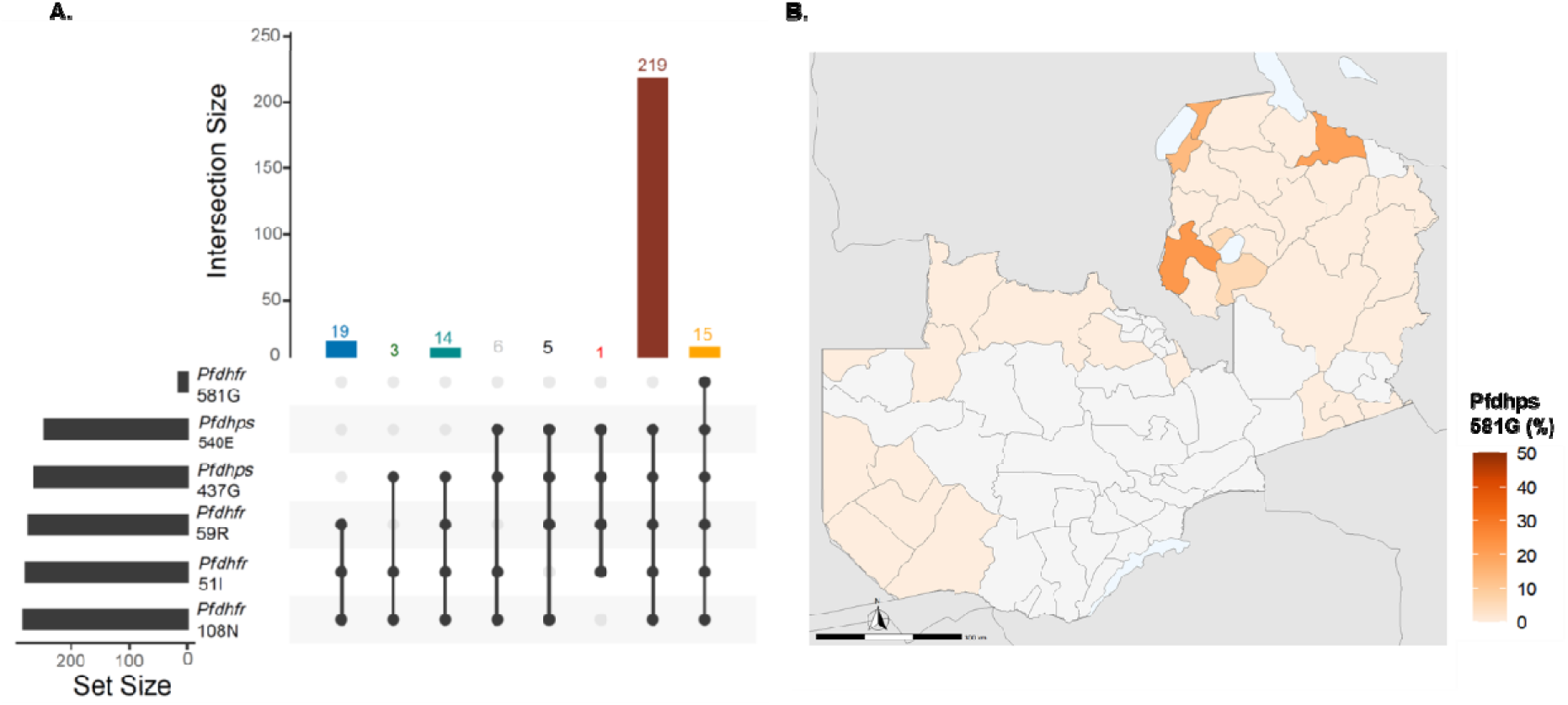
Cumulative prevalence of *Pfdhfr-dhps* mutations (A) and spatial prevalence of *Pfdhps* A581G (B) across Zambia. Color code heat map shows prevalence at the district-level, while white indicates districts where no samples were available.

### Partial artemisinin resistance

A total of 29 mutations were identified across the entire *kelch13* gene, of which 8 were located inside the propeller domain (**Figure 4**). Of these, three mutations (P441**L**: n**=**1 from Western Province, V568**M**: n=2 from Luapula Province and R622**T**: n**=**1 from Western Province) were WHO-validated mutations associated with partial artemisinin resistance. Apart from the K189**T** mutation that was found in 20.6% (58/282) of sequenced samples and was reported in other clinical studies, all other mutations were found at low frequency (<2.5%). The A578**S** mutation, which was identified in seven specimens (2.5%), has been commonly reported in other African countries, although this mutation is not associated with ART resistance in vitro and/or delayed parasite clearance. Overall, 35.1% (99/282) of sequenced samples carried one or more *Pfkelch13* mutations with variable prevalence at the provincial level (**Table S3**), suggesting high polymorphism in the *Pfkelch13* gene due to increased ACT pressure in Zambia.

**Figure 4.**
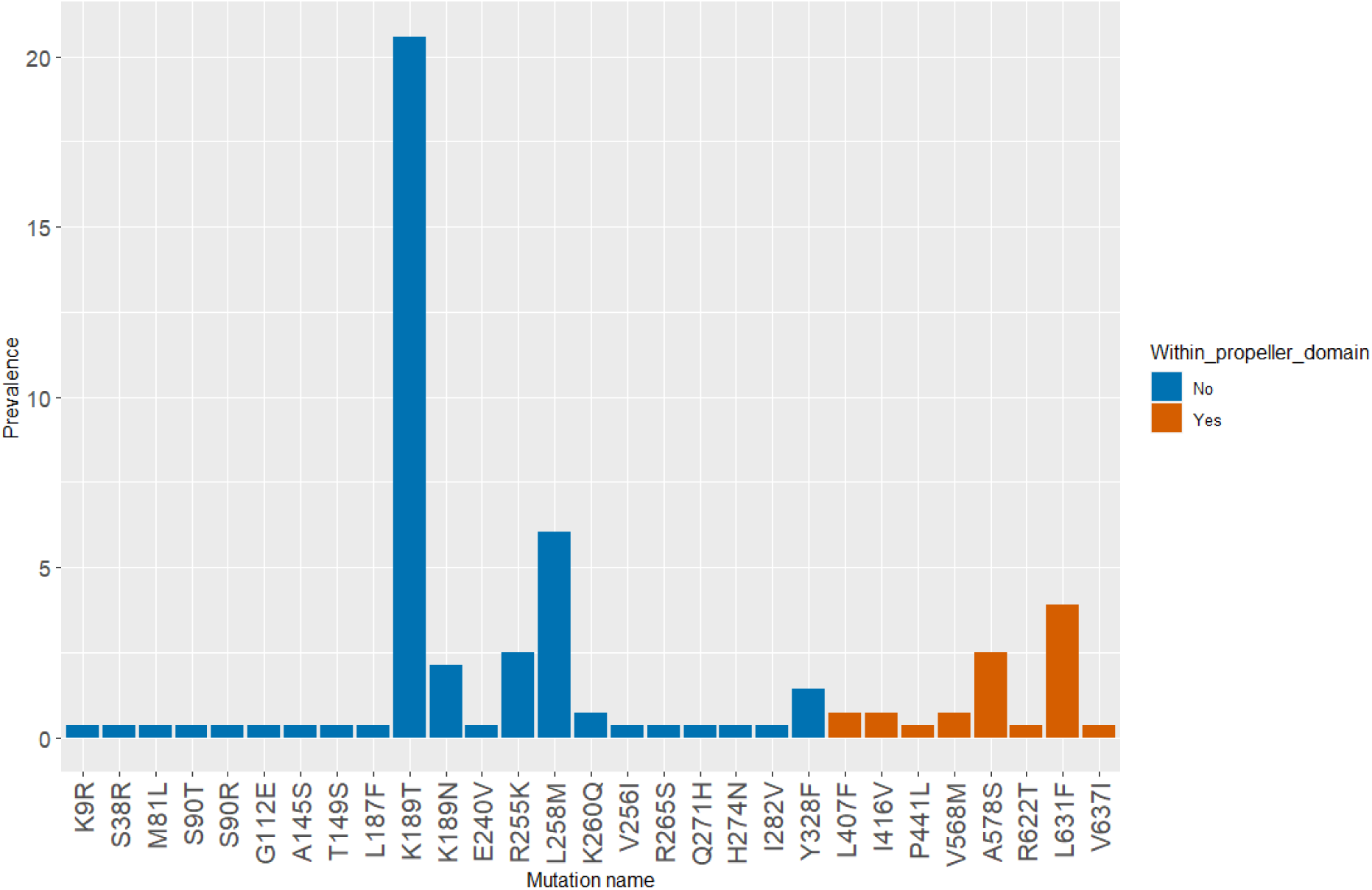
Non-synonymous mutations identified across the *kelch13* gene in Zambia. Colors indicate mutations as outside (blue) or within (orange) the propeller domain.

### Lumefantrine resistance

*P. falciparum* parasite harboring N86 (wild-type), 184F (mutant), and D1246 (wild-type) genotypes in the multi-drug resistance gene 1 (*Pfmdr1*), are associated with decreased sensitivity to lumefantrine. Our analysis revealed that 99.3% (280/282) of the samples carried N86, 55.0% (155/282) of the samples carried 184F, and 98.2% (277/282) carried D1246 (**Figure 5A**). Overall, 53.9% (152/282) of samples carried N86/184**F**/D1246 (N**F**D) genotype with marked spatial variation across districts (**Figure 5B**).

**Figure 5.**
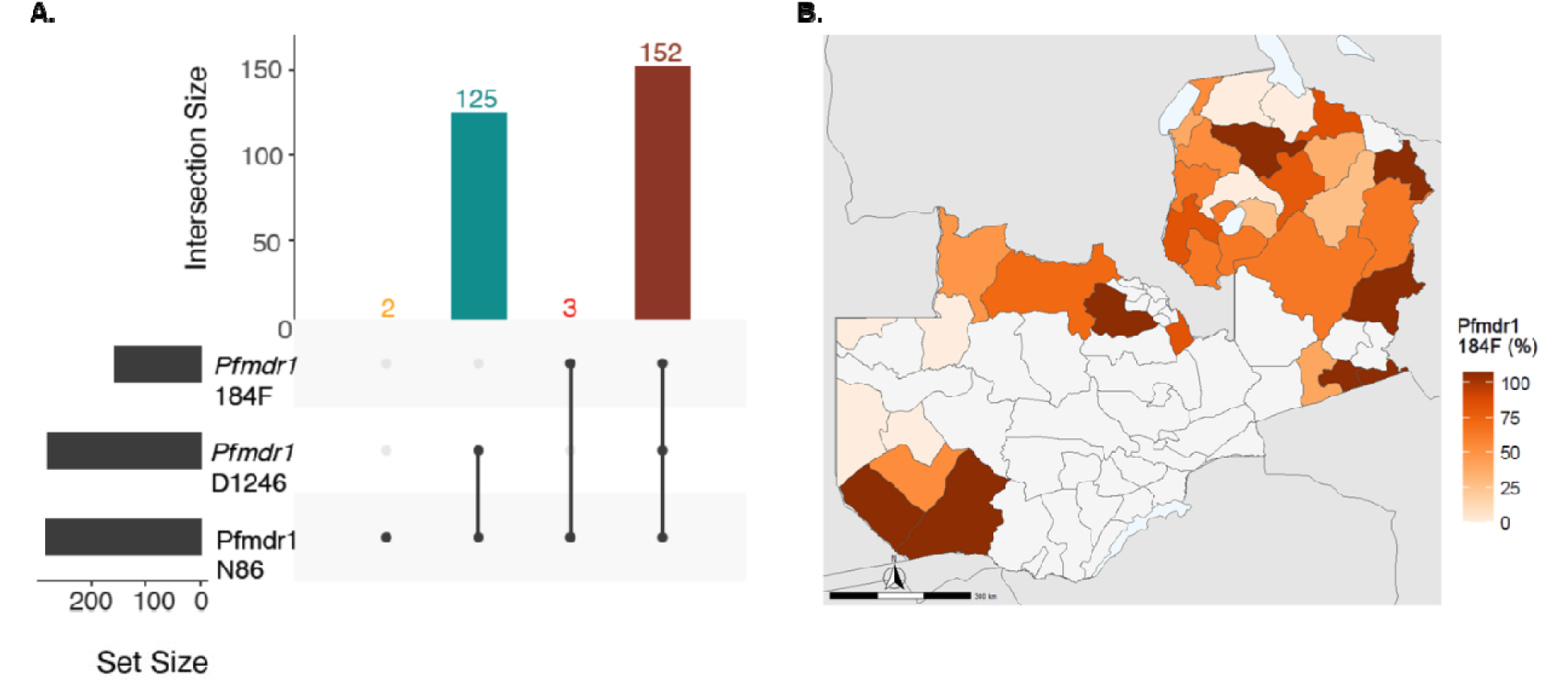
Prevalence of *Pfmdr1* mutations associated with decreased sensitivity to lumefantrine. **A)** Cumulative prevalence of *Pfmdr1* haplotypes, **B)** Spatial prevalence of th *Pfmdr1* 184**F** mutation. Color code heat map shows the prevalence at the district level, while white indicates districts where no samples were available.

### Additional ART resistance-associated mutations

Several mutations in the *Pfatp6* gene, a sarcoplasmic and endoplasmic reticulum Ca^2+^ ATPase (SERCA)-type protein previously associated with artemisinin resistance were identified (**Table S2**). Most mutations occurred at low frequency except for *Pfatp6* N569**K** (54.6%), E431**K** (32.3%), L402**V** (9.6%), and H243**Y** (3.5%). Of the mutations previously shown to mediate resistance, E431**K** (32.3%) and A623**E** (0.35%) were identified, while S769**N** and L263**E** were not reported (**Figure S4**). No other mutations were identified in *Pfarps10*, *Pffd*, *Pfmdr2*, *Pfpib*, *Pfpp*, *Pfap2mu*, *Pfubp1*, and *Pfcrt*, that could potentially augment artemisinin resistance. Several mutations of variable but generally low frequency (**Table S2**) and variable spatial distribution (**Table S3**) were identified in these genes (**Figure S5**).

## Discussion

Intense control activities, especially the continuous use of effective antimalarial drugs, apply significant selective pressure on the malaria parasite population. As transmission declines, most infected individuals carry single clones promoting a higher rate of inbreeding^37^ that favors the spread of drug resistance phenotypes when they arise.^38,39^ The independent emergence or spread of artemisinin resistance in Zambia due to drug selection pressure could significantly increase the malaria burden leading to malaria resurgence and increased morbidity and mortality.^40^ Close monitoring of the efficacy of available antimalarial drugs and improved surveillance for resistance mutations will be key to inform control strategies such that rapid action can be taken to mitigate the impact and slow or prevent the spread of drug resistant parasites.^22,23^

This nationally representative spatial analysis of antimalarial drug resistance mutations supports several conclusions that together strongly suggest that individual genes are under markedly different selection pressures. Firstly, there was a noticeable lack of mutations in *Pfcrt* codons 72-76 and a low number of polymorphisms (only seven unique NS mutations) across the gene. Zambia withdrew CQ treatment and therefore drug selective pressure on the *Pfcrt* gene in 2003 allowing reversion to the wild-type. While other studies have previously identified this reversion,^35,41^ the selection patterns in Zambia lacked a commonly selected region on chromosome 7 (*Pfcrt*), contrasting with parasite populations from other regions. Similarly, we did not observe selection signatures in *Pfaat1*, the second important transporter gene for chloroquine resistance.^42^ This is consistent with other African countries where CQ withdrawal resulted in declines in CQ resistance alleles and reductions in CQ median IC_50_ values.^43,44^ While this finding is encouraging in terms of the potential to reintroduce CQ as part of the chemotherapeutic arsenal in Zambia, preferably as a combination therapy, however additional phenotype-genotype association studies would be needed to confirm the susceptibility of this wild-type strain to CQ.

In contrast to CQ, SP resistance markers were highly prevalent throughout Zambia, suggesting strong selective pressure is maintained through national IPTp implementation and high private sector SP utilization without prescription for self-medication of suspected malaria.^45^ Pyrimethamine associated resistance in *Pfdhfr* was very high, with 95% of samples having triple mutants (IRN) and codon S108N (99.6%) approaching fixation with negligible spatial variation. A similar picture was observed for sulfadoxine associated *Pfdhps* mutations, with 84% double mutants (codons A437**G** and K540**E**). Overall, 82.9% of samples were *Pfdhfr* and *Pfdhps* quintuple mutants *Pfdhfr-dhps* (IRNGE) correlating with full genotypic SP resistance and expected treatment failure. Furthermore, with a concentration in Luapula and Northern Provinces, 5.3% (15/282) of samples also carried the *Pfdhps* A581**G** mutation in addition to the *Pfdhfr-dhps* IRNGE background. This genotype confers extreme SP resistance and is of concern for SP efficacy in Zambia. While some variation between this study and historical data^29,46^ may be explained by study differences (subject selection, sites, implementation period) and data type (PCR genotyping vs. WGS), overall a marked increase in SP genotypic resistance has occurred. Our findings are consistent with our previous evidence of positive selection for SP markers,^31^ as well as with other African countries where *Pfdhfr*–*Pfdhps* quintuple mutant prevalence is high, while the sextuple mutant remains rare,^45,47^ albeit with high spatial heterogeneity.^4,48^ While overall SP resistance is clearly high in Zambia, two mutations (*Pfdhfr* I164**L**^49^ and *Pfdhps* A613**S**/**T**^47,50^) that confer even higher SP resistance were not identified in Zambia.

The high prevalence of SP resistance (>90%) are in children younger than five years) indicates a strong selective pressure has been applied to these genes, even though SP is primarily only used for IPTp. The WHO recommends that countries withdraw SP for IPTp use when the prevalence of *Pfdhps* K540**E** is >95% and *Pfdhps* A581**G** is >10%.^11^ At 87.6% and 5% respectively, Zambia as a country remains below these thresholds, although some districts e.g., Nchelenge and Mansa in Luapula Province, did exceed them. Based on these findings, SP should continue to be used for IPTp. In contrast, the WHO threshold for SP-based IPTi withdrawal is when K540**E** is >50% and thus, SP would not be recommended to be used for IPTi in Zambia at this time.

Until novel therapies are developed, maintaining the efficacy of ART based treatments is fundamental to global control and elimination efforts. In Zambia, AL has been used as a first line combination antimalarial treatment for uncomplicated *P. falciparum* malaria since 2002.^51^ Considering the historical use and importance of ART to malaria control in Zambia, increased polymorphisms in the *kelch13* gene and the identification three WHO-validated mutations associated with partial artemisinin resistance suggest an early signal of partial artemisinin resistance in Zambia. Close monitoring of local emerging or spreading *kelch13* mutations (R561**H**, A675**V** and C469**Y**)^52–54^ that were recently reported from East Africa and confer partial artemisinin resistance is warranted. While not unexpected, it was also encouraging to note that no mutations (*Pfcrt* I356**T**, *Pffd* D193**Y**, *Pfmdr2* T484**I**, *Pfap2mu* S160**N** and *Pfubp1* E1528**D)** associated with ART resistance in Southeast Asia^55^ were identified. Nevertheless, considering the variation between Asian and African parasite populations,^56^ it is possible that other Africa-specific mutations may augment ART resistance. Similarly, we must continue to track all mutations in any key genes, irrespective of their genotypic resistance status. For example, 26 *Pfkelch13* mutations were identified, including one (A578**S**) that has been commonly reported in Africa,^2,29^ the implications of which remains unclear. Unfortunately, while ART appears to be efficacious, the main partner drug lumefantrine does not fare as well, with all but two specimens containing one or more key mutations in *Pfmdr1*. In fact, more than 50% of all specimens carried mdr1 (N**F**D) haplotype. This confirms results provided by other studies performed in the Southern and Western Provinces of Zambia^2,29,57^ but, as with SP, the trend is that genotypic resistance is increasing. While this may not correlate with ACT clinical treatment failure with AL, it does potentially remove the partner drug from the combination therapy leaving ART exposed as monotherapy. Such an environment would be primed to enable rapid selection and spread of ART resistance irrespective of resistance evolving independently or through an introduction event into Zambia.

In summary, this study support two worrying and two encouraging conclusions with respect to antimalarial drug resistance. Firstly, Zambia has very high, and for some loci almost fixed, resistance to SP. While still under WHO recommended limits, this warrants further SP efficacy studies in pregnancy to assess the drugs’ ability to reduce deleterious maternal and birth outcomes, especially in Luapula and Northern Provinces where WHO frequency thresholds were crossed. Secondly, there are also very high levels of genotypic resistance to lumefantrine, the main ART partner ACT drug used in Zambia. While therapeutic efficacy studies have not identified significant treatment failure several years after these samples were collected, it may be prudent to switch to an alternative ACT in the near future, or at least prepare for a switch should treatment failures occur. Finally, there is some encouraging data, namely that CQ sensitivity has been restored and there is no evidence of ART resistance in Zambia. Together these findings along with the recent evidence of strong positive selection signatures genes involved in sulfadoxine-pyrimethamine and artemisinin combination therapies drug resistance^31^ highlight the need of sustained surveillance of antimalarial drug resistance across the country. Furthermore, this work underlines the utility of high-quality genomic surveillance, which if performed and acted upon, gives every chance of effective malaria treatment continuing for the foreseeable future despite the constant threat of drug resistance. Without surveillance, resistance will only be detected following treatment failure, at which point options to respond will be limited.

## Methods

### Sample selection and whole-genome sequencing

This work is a secondary analysis of a larger parent study focused on understanding malaria transmission across Zambia and establishing a baseline for parasite genetic metrics.^31^ Briefly, whole-genome sequencing was performed on 459 *P. falciparum* PET-PCR positive dried blood spots (DBS) collected from children in all ten provinces of Zambia as part of the 2018 Zambia National Malaria Indicator Survey (MIS).^58^ The samples were processed for genomic DNA extraction as previously described.^31, 59^ We adopted a 4-plex hybrid capture method using SeqCap EZ custom probes^60^ to selectively enrich *P. falciparum* genomes prior to sequencing as previously described.^31^ Genomic libraries and hybridization capture were prepared using a modified Roche/Nimblegen SeqCap EZ Library Short Read protocol and sequenced on Illumina NextSeq 6000 (2 × 101-bp) at Yale Center for Genomic Analysis with a target of 30 million reads per sample. Raw sequence reads are available at the Sequence Read Archive (PRJNA932927).

### Variant identification

We identified *P. falciparum* genomic variation from whole genome sequence data with the pipeline previously described.^31,61^ Briefly, Illumina raw paired-end reads were aligned to the *P. falciparum* 3D7 reference genome with BWA-MEM 0.7.17^62^ and removed using Picard Tools 2.20.8. For this study, variant calling was performed only on samples with >30% *P. falciparum* 3D7 reference genome with >5X coverage, resulting in a total of 282 *P. falciparum* samples. Variants were called using GATK v4.1.4.121 following best practices (https://software.broadinstitute.org/gatk/best-practices). We used GATK HaplotypeCaller in GVCF mode to call single-sample variants (ploidy 2 and standard-min-confidence-threshold for calling = 30), followed by GenotypeGVCFs to genotype the parasites. Prior to variant filtering, we scored 1,219,517 SNPs with a VQSLOD >0 across the 282 genomes. The VCF was functionally annotated with SnpEff v4.3 (build 2017-11-24 10:18). Variants removed included those located in telomeric and hypervariable regions (ftp://ngs.sanger.ac.uk/production/malaria/pf-crosses/1.0/regions-20130225.onebased.txt), SNPs with >20% missingness, and minor allele frequency (MAF) >0.02, leaving a total of 27,163 high quality biallelic SNPs.

### Mining drug resistance loci and estimating mutation prevalence

SNPs located in 13 genes associated with antimalarial drug resistance (**Table S1**) were identified using the *VariantAnnotation* R package. All non-synonymous mutations with high read coverage (≥30x) were identified and classified as mutant (heterozygous or homozygous mutant) or wild-type (homozygous reference). The reference allele for *Pfdhps* 437 encodes the mutant allele and was therefore re-coded to A437**G** for clarity as the reference carries a mutant allele unlike all other alleles. The prevalence of each mutation was calculated as (p = m/n*100, where p = prevalence, m = number of infections with mutant alleles, n = number of successfully genotyped infections) using R software version 4.2.0. Mutant combinations were plotted and visualized using the UpSet package in R^63^ and maps were created using the sf package in R.^64^

### Ethical statement

All study participants, and/or their parents or legal guardians, provided written informed consent, and this study was conducted with the approval of the Biomedical Research Ethics Committee from the University of Zambia (Ref 011-02-18) and from the Zambian National Health Research Authority.

## Data availability

The sequence data for the parent project are available in the NCBI Sequence Read Archive, in BioProject PRJNA932927.

## Acknowledgments

The authors are grateful to the Zambian communities, particularly the volunteers and their families, for providing samples during the MIS. We would like to thank the staff of the Zambia National Malaria Elimination Centre for their generous support, especially the field researchers who conducted the nationwide survey.

## Authors’ contributions

G.C., W.J.M and D.J.B. contributed to funding acquisition, project resources and supervision. G.C., A.A.F., W.J.M., and D.J.B., conceived and designed the study. A.A.F., D.J.B. and G.C., coordinated sample selection and curation. M.C.M., B.M., C.M., R.K., M.B.H., B.H., J.M.M. and D.J.B. collected samples and epidemiological data. A.A.F., I.C., and J.D. performed laboratory analysis. A.A.F., D.J.B, G.C. contributed to formal genomic analysis, visualization, interpretation and writing the original draft. All authors contributed to review and editing the manuscript.

## SUPPLEMENTAL MATERIAL

### SUPPLEMENTAL FIGURES

**Figure S1.**
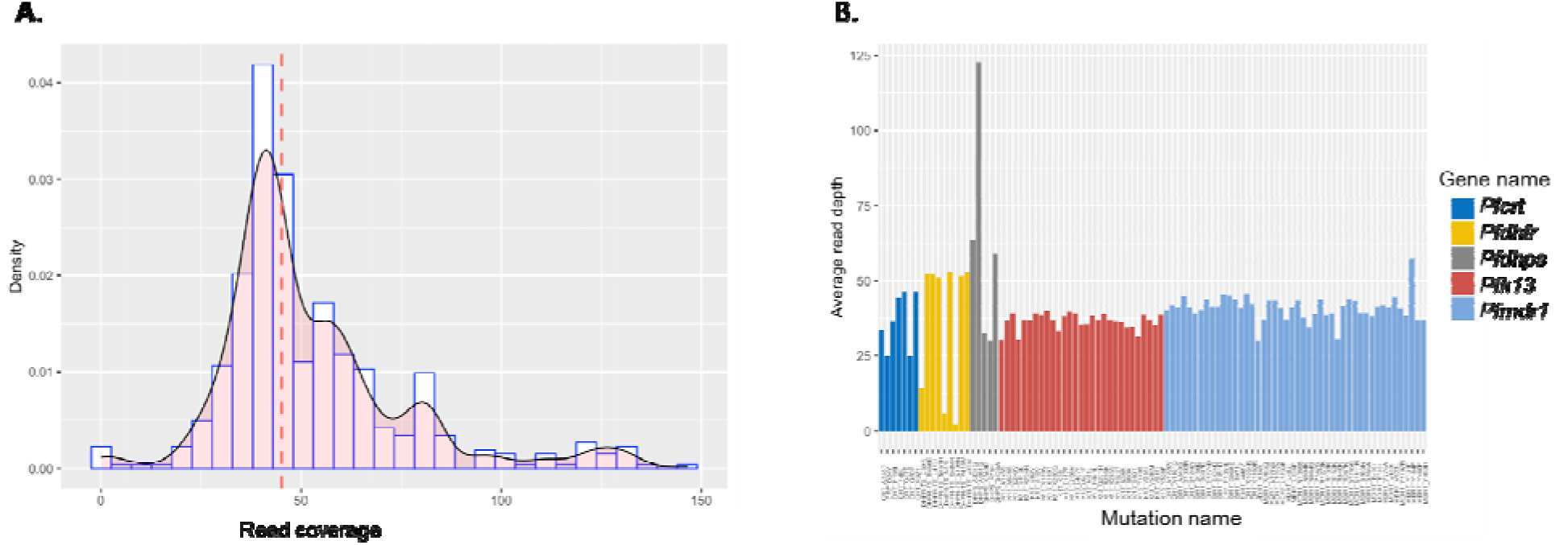
Read coverage per all identified mutations associated with antimalarial drug resistance across 282 samples. **A**) Density plot showing cumulative read depth and dashed red lines indicate median coverage=45X. **B**) Read coverage per mutation for top 5 known antimalarial drug resistance genes.

**Figure S2.**
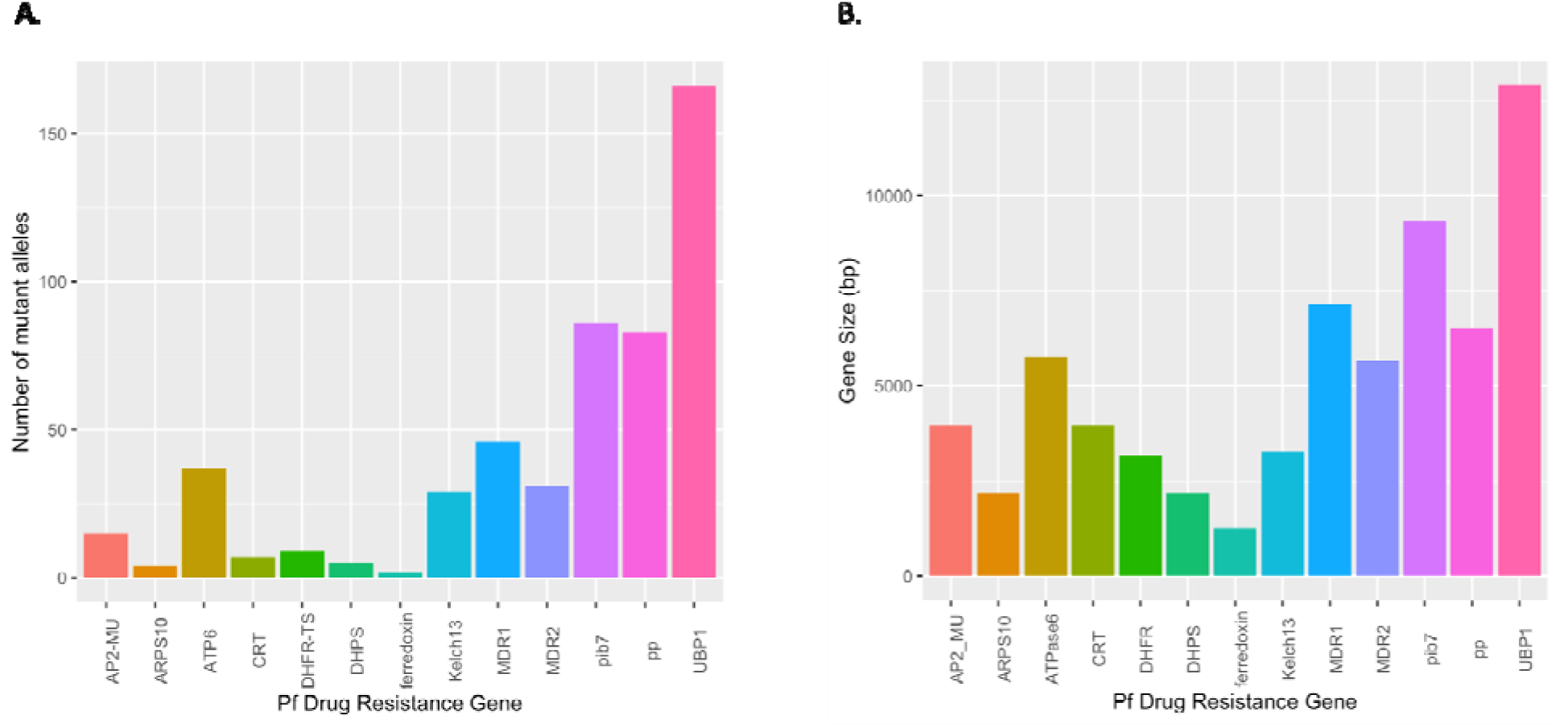
Number of mutations per gene and read depth. **A**) Number of mutations identified across 13 *P. falciparum* genes from 282 samples. **B**) Drug resistance gene size in base pairs.

**Figure S3.**
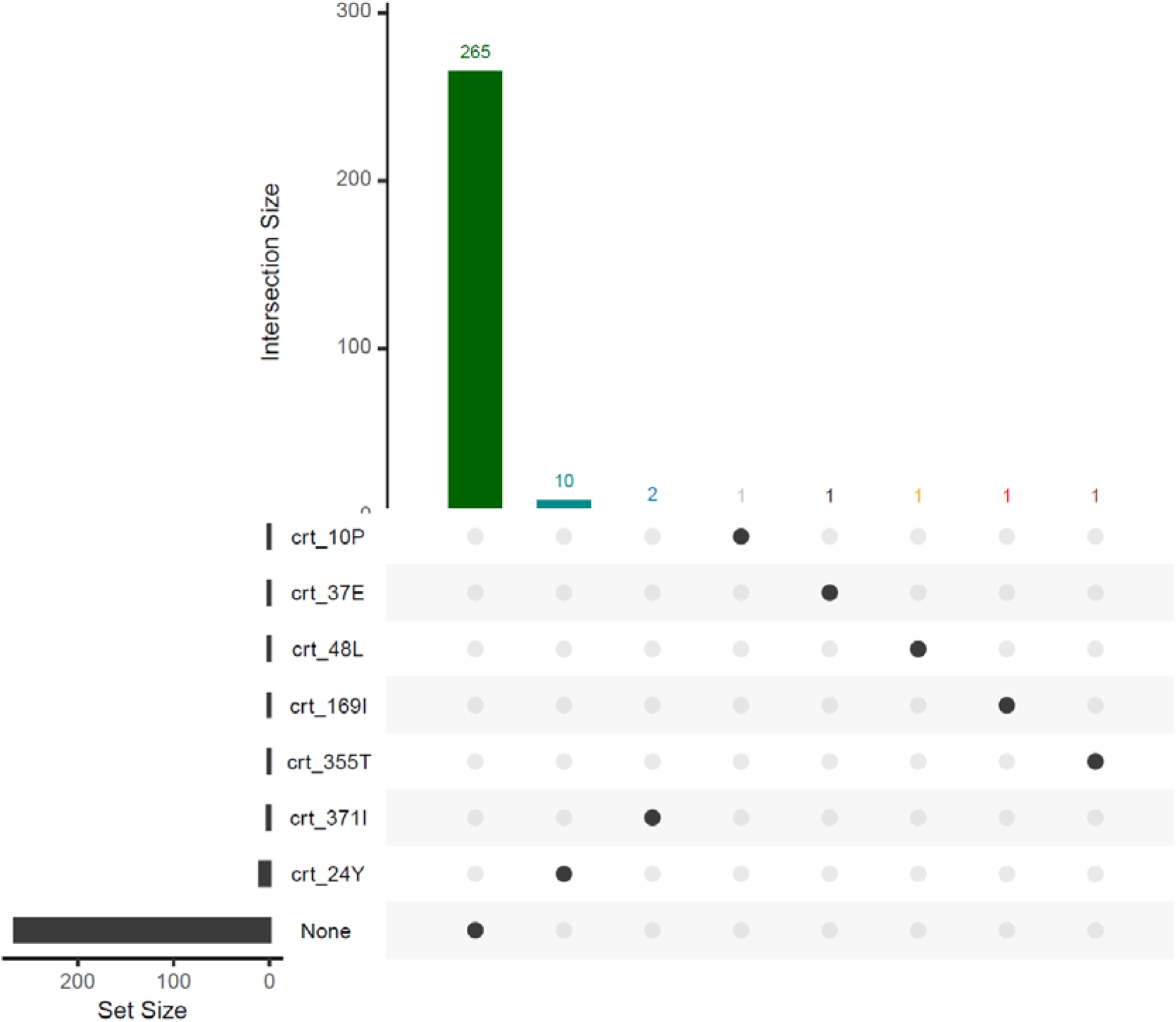
Frequency of mutations in the *Pfcrt* gene. The intersection size bar plots represent the number of parasites carrying different combinations of antimalarial drug resistanc associated mutations in the *Pfcrt* gene. The circles below the bars represent the different combinations of resistant genotypes in individual samples. The set size represents the number of samples in which individual drug resistance-conferring markers were genotyped. Gene position and nucleotide change for each mutation are shown in Table S1. None = samples carrying synonymous mutations or wild-type alleles

**Figure S4.**
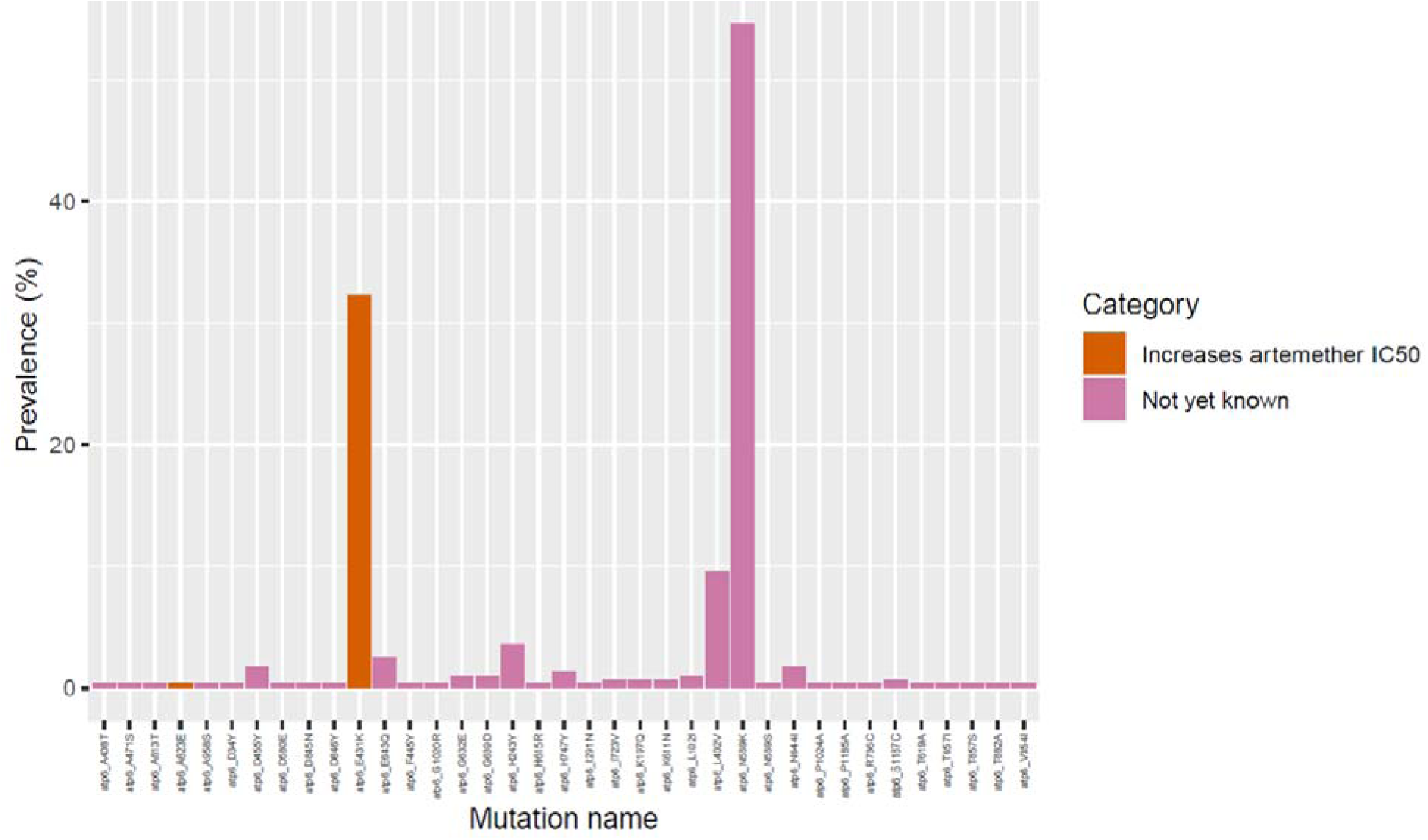
Prevalence of 37 mutations in *Pfatpase6* from 282 Zambian samples ordered by genomic loci. Colours indicate whether the mutations are validated or not as resistance markers for ACT.

**Figure S5.**
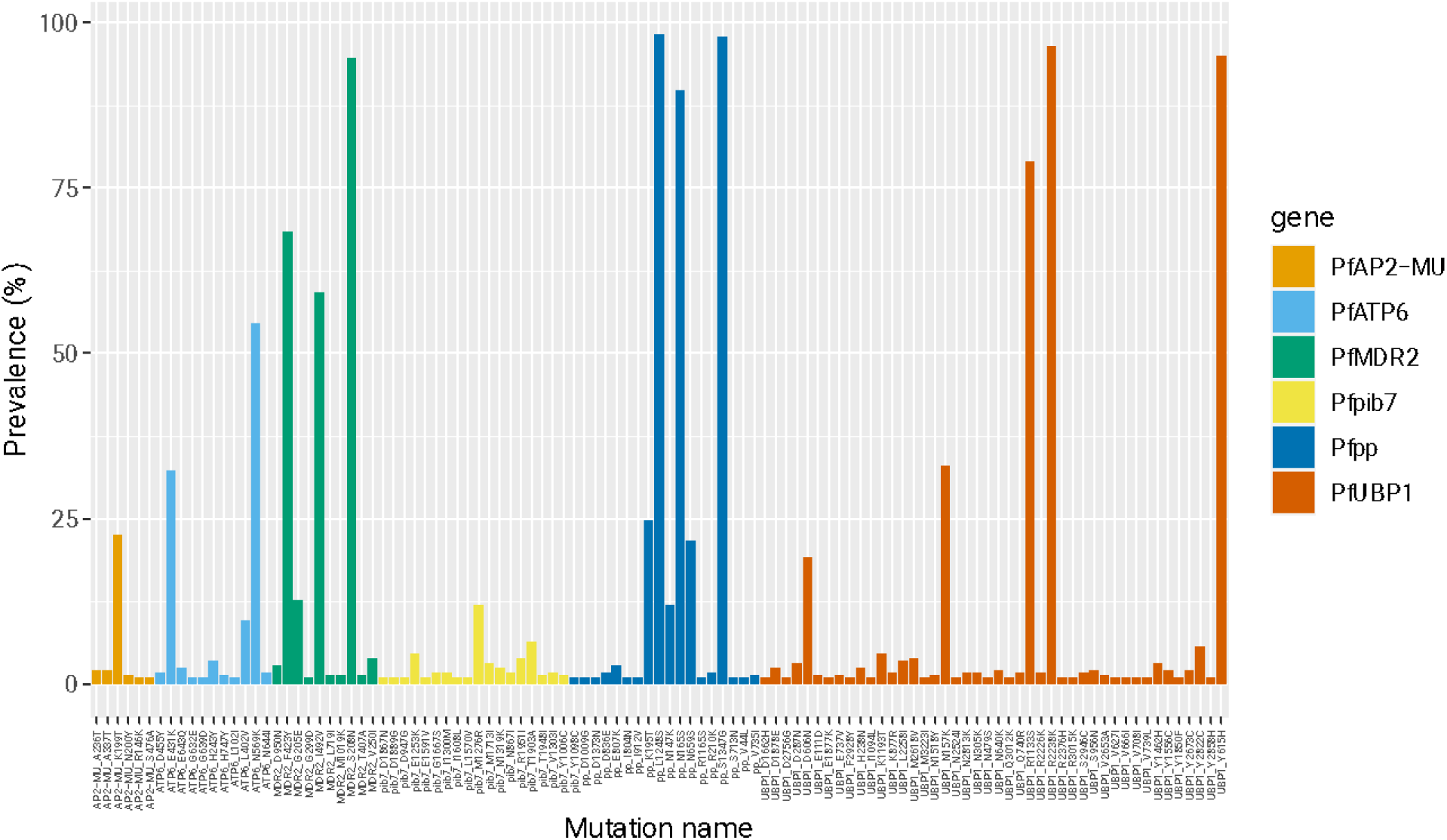
Prevalence of different mutations across genes associated with partial artemisinin resistance other than *Kelch13* gene. We identified several mutations across *PfATPase6* protein, apicoplast ribosomal protein S10 precursor gene (*Pfarps10*), (*Pfmdr*) protein 2 codon, AP-2 complex subunit mu gene (*Pfap2-mu*), ubiquitin carboxyl-terminal hydrolase 1 gene (*Pfubp-1*) and Phosphoinositide-binding protein (*Pfpib7*). No mutation observed in genes: *Pfcrt* I356**T**, *Pffd* D193**Y**, *Pfmdr2* T484I, *Pfap2-mu* S160N/T and *Pfubp-1* E1528D – known background mutations associated with ART in SEA. Also, no mutation was observed in the AP-2 complex subunit mu gene (ap2-mu, S160N/T) and the ubiquitin carboxyl-terminal hydrolase 1 gene (ubp-1, E1528D), mutation that is associated with delayed ACT clearance in Africa.

### SUPPLEMENTAL TABLES

*Supplementary tables are compiled into a single file for ease of viewing*

**Table S1. Description of 13 *P. falciparum* drug resistance genes across successfully sequenced samples and included in this analysis and the drugs with which they are associated.**

See uploaded file.

**Table S2. Sequencing coverage across successfully sequenced samples and cumulative prevalence of all mutations across 13 genes in Zambia.** n = number of samples successfully sequenced samples

See uploaded file.

**Table S3. Prevalence of all mutations across 13 genes per region in Zambia.** n = number of samples successfully sequenced samples

See uploaded file.

